# Multinational Assessment of Absolute Neutrophil Counts and White Blood Cell Counts Among Healthy Duffy Null Adults

**DOI:** 10.1101/2025.03.27.25324712

**Authors:** Stephen P. Hibbs, Israel Chipare, Amr J Halawani, Sophie E Legge, Geoffrey Fell, Daniel Dees, Abdulrahman A Alhamzi, Edwig Shingenge, Mohammed J Alabdly, Hilary T Charuma, Mohammed A Nushaily, Judith M Sinvula, Menelik Russo, Michelle Sholzberg, Sara Paparini, Vanessa Apea, Maureen Achebe, Nancy Berliner, Lauren E. Merz

**Affiliations:** Wolfson Institute for Population Health, Queen Mary University of London, London, United Kingdom; Blood Transfusion Service of Namibia, Windhoek, Namibia; Department of Clinical Laboratory Sciences, Faculty of Applied Medical Sciences, Umm Al-Qura University, Makkah, Saudi Arabia; Centre for Neuropsychiatric Genetics and Genomics, Division of Psychological Medicine and Clinical Neurosciences, Cardiff University, Cardiff, United Kingdom; Data Science, Dana-Farber Cancer Institute, Boston; Department of Pathology, Brigham and Women’s Hospital, Boston, MA, USA; Department of Blood Bank, King Fahad Central Hospital, Jazan, Saudi Arabia; Department of Clinical Health Sciences, Namibia University of Science and Technology, Windhoek, Namibia; Department of Laboratory Medicine and Pathobiology, University of Toronto, Toronto, Canada; Department of Medicine, University of Toronto, Toronto, Canada; Department of Medicine, and Department of Laboratory Medicine and Pathobiology, St. Michael’s Hospital, Li Ka Shing Knowledge Institute, Toronto, Canada; Division of Hematology, Department of Medicine, Brigham and Women’s Hospital, Boston, MA; Division of Hematology/Oncology, Mass General Brigham, Boston, MA; Department of Medicine, Harvard Medical School, Boston, MA; Department of Medical Oncology, Dana-Farber Cancer Institute, Boston, MA

## Abstract

**Background:** Laboratory reference intervals must reflect population diversity for accurate medical decisions. The Duffy null variant lowers absolute neutrophil counts (ANC), but existing dedicated reference intervals are based on a single African American cohort. The impact across other ethnic groups and regions remains unclear, and no white blood cell count (WBC) intervals exist for Duffy null individuals. This study aimed to establish and compare Duffy null ANC and WBC reference intervals across four continents.

**Methods:** A cross-sectional study was conducted assessing healthy Duffy null individuals from dedicated cohorts (blood donors in Namibia and Saudi Arabia, and primary care patients in the USA) and biobanks (participants from the UK and USA). Reference intervals were determined using Clinical & Laboratory Standards Institute guidelines.

**Results:** Among 7,872 participants (392 from dedicated cohorts, 7,480 from biobanks), novel ANC and WBC reference intervals were established: Namibia (820–6,370/μL; 2.51–9.85× 10^9^/L), Saudi Arabia (1,090–5,100/μL; 3.71–9.95× 10^9^/L), and the USA (1,210–5,390/μL; 3.00-9.66× 10^9^/L), with no significant differences between cohorts. Institutional reference intervals misclassified 27.9% (Namibia), 50.7% (Saudi Arabia), and 21.7% (USA) as neutropenic. Biobank analyses confirmed a significantly lower ANC in Duffy null individuals compared to Duffy non-null (p<0.0001) with no significant difference in ANC between Black and non-Black Duffy null participants.

**Conclusions:** Duffy null individuals consistently exhibit lower ANC and WBC across ethnic groups and regions. Current reference intervals overlook this variation, risking misdiagnosis and health inequities. Implementing Duffy-specific reference intervals is essential for equitable and accurate clinical decisions worldwide.

**Key points:**

– Novel ANC and WBC reference intervals for Duffy null adults were established and are consistent across four continents
– Current ANC reference intervals misclassify up to half of Duffy null individuals as neutropenic, contributing to global heath inequities.

## INTRODUCTION

Laboratory reference intervals guide clinicians in distinguishing between normal and abnormal results. These intervals are typically established using samples from approximately 120 healthy individuals within a healthcare system.^1^ Subsequently, other laboratories may adopt these standards using a verification sample of as few as 20 individuals.^1^ However, small reference samples often fail to reflect diversity within communities, potentially compromising clinical decision-making.

An example of this is seen in absolute neutrophil counts (ANC) which show significant global variation. ANC reference intervals in Zimbabwe (939-4252 cells/uL)^2^, Uganda, (900-3900 cells/uL)^3^, and Kenya (1050-4080 cells/uL)^4^ differ substantially from those used in the UK (2000-7000 cells/uL)^5^ and North America (2500-7000 cells/uL).^6^ This variation in ANC is predominantly driven by a single nucleotide polymorphism (SNP) in the promoter region of the *ACKR1* gene which prevents transcription and leads to the erythrocyte Duffy null phenotype [Fy(a−b−)].^7^ Mouse models suggest that this SNP impacts hematopoiesis, producing phenotypically distinct neutrophils that leave the periphery and preferentially localize to the spleen.^8,9^ Despite lower circulating neutrophil counts, individuals with Duffy null status maintain normal total body neutrophil counts, bone marrow cellularity, and response to infection^10,11^. Additionally, Duffy null status confers partial protection against *Plasmodium vivax^7,12^*, explaining its prevalence in certain endemic regions like Sub-Saharan Africa and the Arabian Peninsula.^13^ For example, the Duffy null phenotype is seen in 80-100% of people living in West Africa, 50-60% living in some Middle Eastern regions, and under 1% of individuals of South Asian, East Asian, or European ancestry^13^.

Although lower ANC is observed more often in certain groupings, the biologic driver of neutrophil variation is the Duffy null genotype^14^. One reference interval for Duffy null individuals has been published, derived from 120 self-identified Black or African American adults in Boston.^15^ However, it is uncertain whether Duffy null-specific ANC reference intervals might vary across countries, genetic ancestries, and racial or ethnic identities due to other unidentified genetic or environmental factors. Additionally, there are no published reference intervals for total white blood cell count (WBC). This multinational study aimed to create and compare Duffy null specific ANC and total WBC reference intervals using both data from directly recruited cohorts and biobanks from diverse populations across four continents. We aimed to establish the degree of variation across different geography or ethnic groupings, and to determine how often current regional reference intervals would misclassify individuals with the Duffy null variant.

## METHODS

We identified and created five different datasets, selected for geographical and ancestral diversity in centers where Duffy phenotyping or rs2814778 genotyping was available. Concordance between genotyping and phenotyping is high.^15^ Datasets included three “dedicated cohorts”: new cohorts of healthy blood donors from Namibia and Saudi Arabia and a previously established cohort of healthy primary care patients in the USA^16^. The other two datasets comprised biobank data from the UK Biobank^17^ and the Mass General Brigham (MGB) Biobank^18^. Each cohort used different approaches to ethnic or racial categorization, reflecting regional variation in labelling group identities, which are detailed below. **Figure 1** outlines the definitions and differences between the terms race, ethnicity, and genetic ancestry.^7^

**Figure 1:**
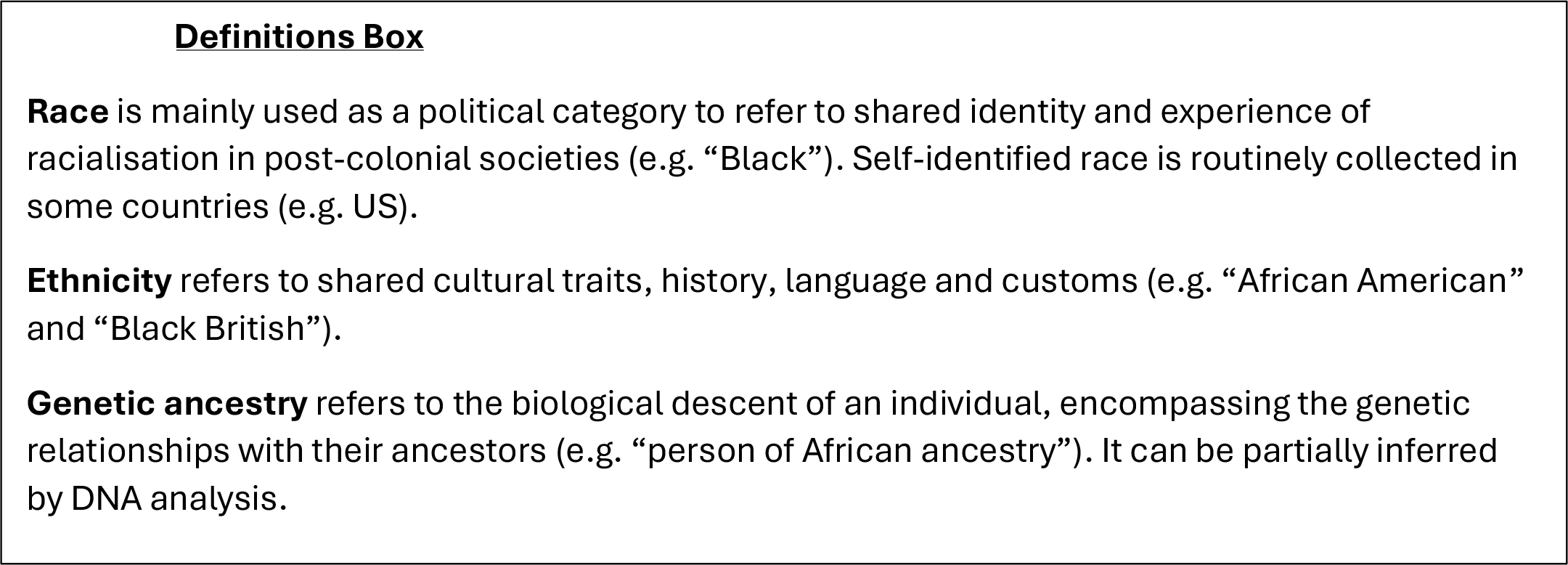
Race, Ethnicity, and Genetic Ancestry Definitions Box. Race, ethnicity, and genetic ancestry are distinct concepts, but are often used interchangeably in medical literature. While no firm consensus exists on the use of these terms, we define and apply them in these ways for the purposes of this study.

### Namibian Blood Donors

Healthy volunteer blood donors attending Blood Transfusion Service of Namibia donor clinics in Windhoek, Khomas Region, were enrolled between September 2023 to December 2023. Donor eligibility criteria are included in **Supplementary Methods**. Donors self-identified as White (predominantly European ancestry), Black (predominantly African ancestry), or Coloured (mixed African, European, and/or Asian ancestry)^19^. Duffy serotyping was performed using Anti-Fya/Fyb IgG antisera by tube testing Rapid Labs, Essex, United Kingdom), and complete blood counts with differential (CBC) were measured with a Sysmex XN-450 analyzer (Sysmex, Kobe, Japan). Ethical approval was obtained from the Ministry of Health and Social Services, Namibia University of Science and Technology Ethical Committee, and the Blood Transfusion Service of Namibia, with written donor consent to collect research samples.

### Saudi Arabian Blood Donors

Healthy volunteer blood donors were enrolled at King Fahad Central Hospital in Jazan Province between December 2023 and January 2024. Donor eligibility criteria are included in **Supplementary Methods**. Duffy serotyping was performed by gel card technology using ID-Card Fya/Fyb and ID-Anti-Fya/Fyb antibodies (DiaMed GmbH, Cressier, Switzerland), and CBC were measured with a Sysmex XN-10 analyzer (Sysmex, Kobe, Japan). Ethical approval was obtained from the Jazan Health Ethics Committee, and written donor consent was obtained.

### USA Primary Care Patients

Patients attending a primary care clinic in Boston, USA for a non-urgent care visit who self-identified as Black or African American were enrolled. Patients with any medication or condition that could impact ANC were excluded (detailed in **Supplementary Methods, eTable 1**). Verbal consent was obtained. CBC was performed on a Sysmex XN-9000 analyzer (Sysmex, Lincolnshire, IL). Duffy phenotyping for Fya and Fyb was performed by tube testing using serologic reagents from Bio-Rad (Hercules, CA) and Quotient (Newtown, PA), respectively. Genotyping was performed using the PreciseType HEA Molecular BeadChip Test (Immucor, Norcross, GA). Local institutional review board approval was obtained.

### UK Biobank Participants

UK Biobank is a biomedical database of approximately 500,000 individuals from across the UK aged 40 to 69 years at recruitment (recruited between 2006 and 2010). Biobank participants were asked to self-report their ethnicity as White British/Irish, Asian, Black African/Caribbean, Chinese, Mixed, Other ethnic group, or prefer not to answer. UK Biobank participants who had both genotyping of the rs2814778 variant and an ANC measurement (field ID: 30140) available were included. Participants were excluded if they had ICD-10 diagnoses associated with alterations in normal neutrophil (detailed in **Supplementary Methods, eTable 2**). All participants provided written informed consent, and ethical approval was granted by the North-West Multi-Centre Ethics Committee. This study was conducted under UK Biobank project number 13310.

### MGB Biobank Participants

Genomic data and health information were obtained from the Mass General Brigham Biobank, a biorepository of consented patient samples at Mass General Brigham in the USA. Participants were included if they had genotyping of the rs2814778 variant and at least one ANC measurement available in the electronic health record (EHR). Subjects with computed phenotypes consistent with HIV infection, systemic autoimmune disease, rheumatoid arthritis, noninfective inflammatory bowel disease, celiac disease, hepatitis C, myeloproliferative disease, or personal history of malignancy were excluded. Laboratory data was obtained through linkage with electronic medical records rather than a dedicated research visit, leaving data susceptible to influence from acute illness. Thus, ANC values ≥20,000/uL were excluded as these were unlikely to represent states of health. The first ANC value available in the electronic medical record was used in the analysis. Local IRB approval was obtained (2024P001516).

### Statistical Analysis

Statistical analysis of the MGB Biobank and dedicated cohorts was conducted in R version 4.2.2 and UK Biobank conducted with R version 4.2.1. Statistical significance was declared at the 0.05 type-I error level.

#### Dedicated cohort analysis

All continuous variables were summarized both parametrically by mean and standard deviation and non-parametrically by median and quantiles. Categorical variables were summarized by counts and percentages. The primary analysis focused on associations with ANC levels, with WBC associations as a secondary objective. Given known differences in ANC by sex, we also tested if differences in ANC and WBC between cohorts were explained by differences in sex distributions^20^.

Before significance testing, both ANC and WBC distributions were found to be non-normal as determined by the Shapiro-Wilks test for normality. All omnibus comparisons of differences across distributions were assessed using the Kolmogorov-Smirnov test when more than two groups were being compared. For all significant omnibus tests, pairwise Wilcoxon tests were then estimated and multiple comparisons were adjusted using Holm’s method. If only two groups were under consideration, the Wilcoxon rank sum test was used.

Following the recommended approach in Clinical & Laboratory Standards Institute (CLSI) EP28-A3c guidelines^1^, EP Evaluator version 12.0.0.11 (Data Innovations) software was used to establish a 95% reference interval by a nonparametric percentile method. To assess the upper and lower bounds of the central 95% ANC and WBC values for the three dedicated cohorts combined, the bootstrap method was employed. We simulated 15,000 datasets of 392 subjects from the USA, Namibia and Saudi Arabia cohorts by sampling with replacement to estimate the 2.5% and 97.5% quantiles. With the bootstrapped standard error we estimated the 95% Gaussian confidence interval around each quantile.

#### Biobank analysis

UK and MGB Biobank analyses grouped rs2814778 genotypes as CC vs. CT/TT and calculated the central 95% ANC distribution for each group. The association between absolute neutrophil count and Duffy-null genotype status (TT vs. TC/CC for rs2814778) was tested via multivariable linear regression adjusted for age and sex in UK Biobank and MGB Biobanks. The association between ethnicity status (Black vs other ethnicities), age and sex was tested via multivariable linear regression in biobank participants with the Duffy-null genotype.

## RESULTS

### Participant demographic information

In total, 122 Duffy null donors in Namibia, 150 Duffy null donors in Saudi Arabia, and 120 healthy Duffy null primary care patients in Boston, USA were recruited. USA participants self-identified as Black or African American and participants in the Saudi Arabian data self-identified as Saudi Arabs. In the Namibia cohort, 87.7% (n=107) self-identified as Black, 6.6% (n=8) as Coloured, and 5.7% (n=7) as White. Demographic data is shown in **Table 1**.

**Table 1:** Demographics, Absolute Neutrophil Count (ANC), and White Blood Cell Count (WBC) of Duffy Null Individuals. Demographics, ANC, and WBC distributions, including the lower limit of normal (2.5th percentile) and upper limit of normal (97.5th percentile) are reported for dedicated cohorts from Namibia, Saudi Arabia, and the USA. Additionally, demographics and ANC distributions, along with their respective lower (2.5th percentile) and upper (97.5th percentile) limits of normal, are reported for the UK Biobank and MGB Biobank.

|  |  | Namibia (n=122) | Saudi Arabia (n=150) | USA (n=120) | MGB Biobank (n=1,563) | UK Biobank (n=5,917) |
| --- | --- | --- | --- | --- | --- | --- |
| **Demographics** | Age, median (25<sup>th</sup> & 75<sup>th</sup> quartiles), *years* | 28 (22-37) | 29.5 (24-38) | 51 (43-61) | 54 (40-66) | 50 (45-56) |
|  | Male sex, %, (n) | 38.5% (47) | 98.7% (148) | 26.7% (32) | 38.6% (602) | 43.6% (2582) |
| **ANC** | Median (25<sup>th</sup> & 75<sup>th</sup>), *cells/uL* | 2,470<br>(1,878-3,445) | 2,475<br>(1,853-3,047) | 2,820<br>(2,090-3,473) | 3,260<br>(2,390-4,654) | 2,640<br>(2100-3300) |
|  | Mean (range), *cells/uL* | 2,822<br>(550-7230) | 2,576<br>(930-6,180) | 2,883<br>(1,080-5,950) | 3,928<br>(600-17,900) | 2,794<br>(120-8870) |
|  | Lower limit of normal-2.5% (95% CI), *cells/uL* | 820<br>(550-1,290) | 1,090<br>(930-1,440) | 1,210<br>(1,080-1,390) | 1,190 | 1,300 |
|  | Upper limit of normal- 97.5% (95% CI), *cells/uL* | 6,370<br>(5,320-7,230) | 5,100<br>(4,670-6,180) | 5,390<br>(4,640-5950) | 10,910 | 5,160 |
| **WBC** | Median (25<sup>th</sup> & 75<sup>th</sup>), *cells × 10<sup>9</sup>/L* | 5.54<br>(4.36-6.65) | 5.89<br>(5.23-7.23) | 5.61<br>(4.59-6.39) | - | - |
|  | Mean (range), *cells × 10<sup>9</sup>/L* | 5.66<br>(2.27-11.36) | 6.29<br>(2.60-10.98) | 5.76<br>(2.69-11.89) | - | - |
|  | Lower limit of normal-2.5% (95% CI), *cells × 10<sup>9</sup>/L* | 2.51<br>(2.27-3.21) | 3.71<br>(2.60-4.02) | 3.00<br>(2.69-3.57) | - | - |
|  | Upper limit of normal-97.5% (95% CI), *cells × 10<sup>9</sup>/L* | 9.85<br>(8.77-11.36) | 9.95<br>(9.13-10.98) | 9.66<br>(8.04-11.89) | - | - |

Figure 2. summarizes the number of participants excluded and included within both biobanks with reasons for exclusion. Within the UK Biobank, 487,323 individuals had genetic data available for rs2814778. A total of 1,533 of participants with the Duffy null genotype (rs2814778 CC) and 139,695 participants without the Duffy null genotype (rs2814778 TT, TC, CT) were excluded for having conditions that could impact ANC (eTable 2). Of the 5,917 eligible participants with the Duffy null genotype, 85.3% (n=5050) self-identified their ethnicity as Black African/Caribbean, 0.3% (n=16) as White British/Irish, 11.6% (n=685) as other or mixed ethnicity, and 2.8% (n=165) preferred not to answer or reported not knowing their ethnicity. Demographic data is shown in **Table 1**.

**Figure 2:**
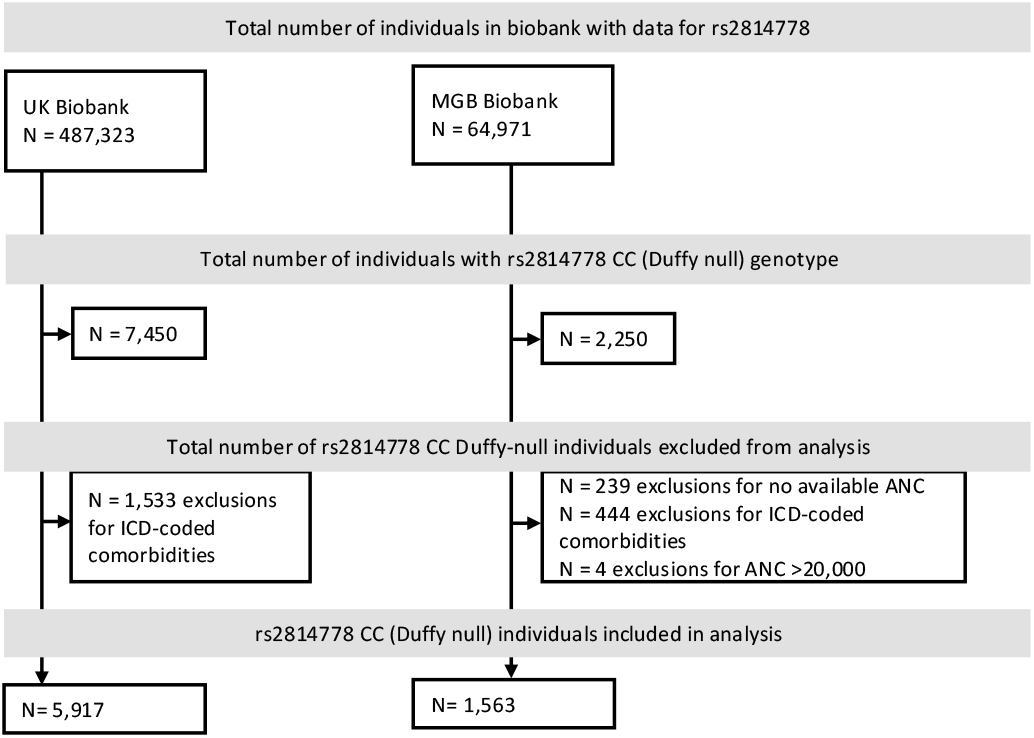
Biobank screening, eligibility, and exclusion process. The figure illustrates the search strategy for identifying individuals in each biobank with available rs2814778 data, the identification of Duffy null (CC) participants, reasons for exclusions, and the final number of individuals included in the analysis.

Within the MGB Biobank, 64,971 individuals had rs2814778 evaluated. Of the 2,250 Duffy null individuals (rs2814778 CC), 2,011 (89.4%) had at least one ANC value available. Of the eligible Duffy null participants, 83.1% (n=1299) self-identified their racial category as Black or African American and 3.4% (n=53) self-identified as White. Demographic data on eligible participants with the Duffy null genotype is shown in **Table 1**. Of the 62,721 samples without the rs2814778 CC variant (Duffy non-null), 28,353 individuals were eligible for analysis after removing those with qualifying phenotypes, those without ANC available, or those with ANC≥20,000 cells/uL. Of the eligible Duffy non-null participants, 84.1% (n=23,838) self-identified as White and 2.6% (n=724) self-identified as Black or African American.

### Absolute Neutrophil Count distributions in Duffy null individuals

ANC distributions in healthy participants with the Duffy null phenotype from dedicated cohorts in Namibia, Saudi Arabia, and USA is shown in **Table 1** and **Figure 3**. The nonparametric central 95% ANC interval indicating the 2.5% and 97.5% values is 820-6,370/uL in Namibia, 1,090-5,100/uL in Saudi Arabia, and 1,210-5,390/uL in the USA^16^. There is an overlap of 95% confidence intervals surrounding the upper and lower limits of normal between all three dedicated cohorts. Bootstrapped analysis of the three dedicated cohorts combined gives a 2.5% and 97.5% ANC value of 1,200/uL (95% CI: 1,060-1,340) and 5,400/uL (95% CI: 4,970-5,810), respectively.

**Figure 3:**
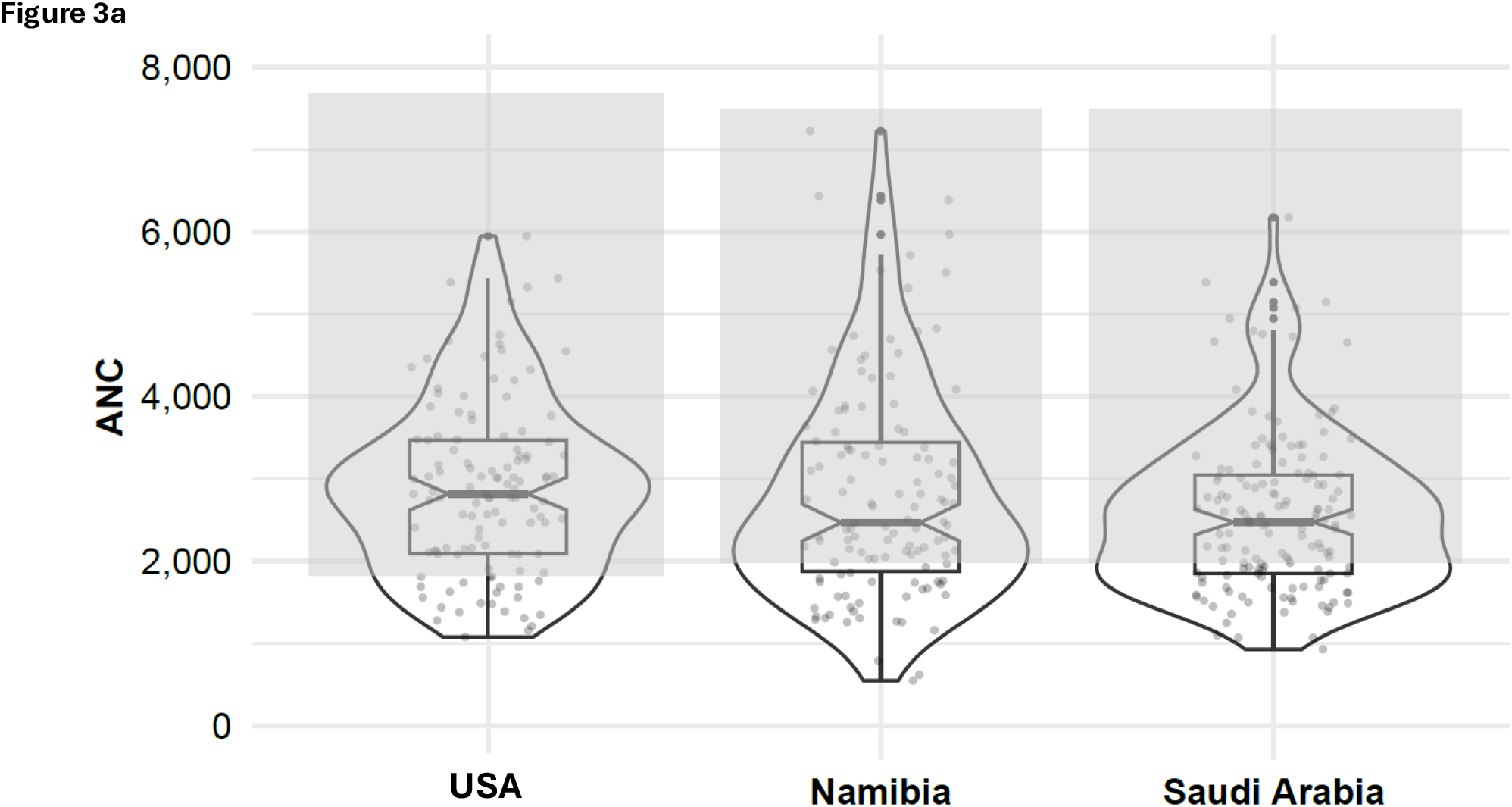
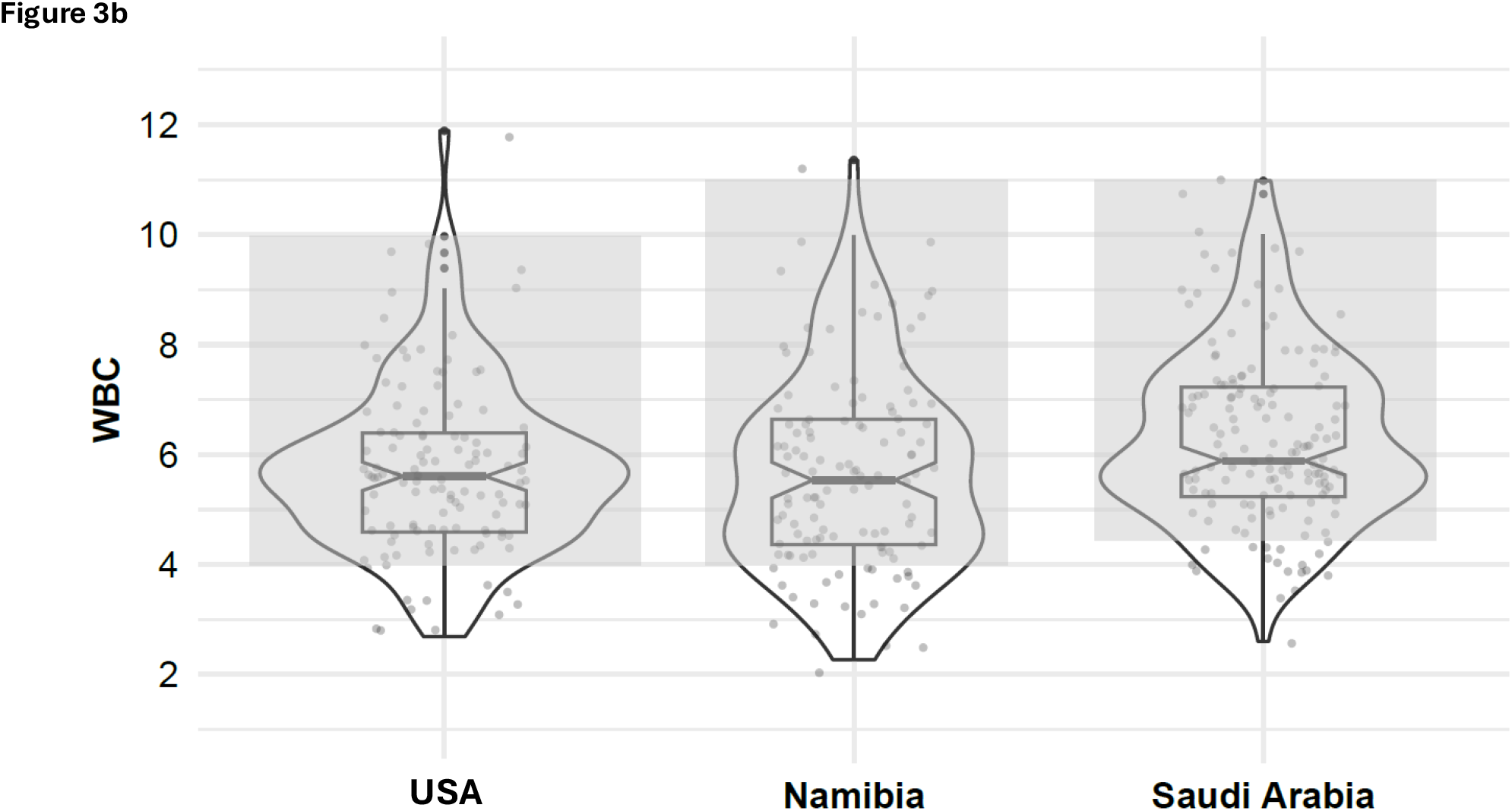
Duffy Null ANC and WBC Distribution with Institutional Reference Intervals. a) ANC distribution and individual values are presented for each of the three dedicated cohorts (USA, Namibia, and Saudi Arabia). Respective institutional reference intervals are indicated by the grey shaded areas: 2000-7500/μL in Namibia; 2500-7500/μL in Jazan, Saudi Arabia; and 1920-7600/μL in Boston, USA. b) WBC distribution and individual values are presented for each of the three dedicated cohorts (USA, Namibia, and Saudi Arabia). Respective institutional reference intervals are indicated by the grey shaded areas: 4.00-11.00× 10^9^ /L in Namibia; 4.50-11.00× 10^9^ /L in Jazan, Saudi Arabia; 4.00-10.00× 10^9^ /L in Boston, USA.

ANC distribution differed across dedicated cohorts (p=0.048). For pairwise comparisons, median ANC did not differ between the USA and Namibia (p = 0.512) nor Namibia and Saudi Arabia (p = 0.512), but did differ between the USA and Saudi Arabia (p = 0.031). However, there is no significant difference in ANC medians between the USA and Saudi Arabia when only male data is analyzed (2475/uL vs 2505/uL, p=0.897).

We assessed the proportion of participants with ANC below their respective institutional lower limit of normal (2000/μL in Namibia; 2500/μL in Jazan, Saudi Arabia; 1920/μL in Boston, USA). In total, 27.9% (n=34) of the Namibia cohort, 50.7% (n=76) of the Saudi Arabia cohort, and 21.7% (n=26) of the USA cohort had an ANC below their respective institutional reference interval.

In the biobanks, data from participants with the Duffy null genotype and Duffy non-null genotypes were analyzed. ANC values from eligible Duffy null individuals in the UK Biobank and MGB Biobank are shown in **Table 1**. The nonparametric central 95% interval for ANC indicating the 2.5% and 97.5% values for Duffy null individuals was 1,300-5,160/uL in the UK Biobank and 1,190-10,910/uL in the MGB Biobank. There was a significant difference in ANC by Duffy genotype (null vs non-null) in both the UK Biobank (ß=−1.41; SE=0.02; 95% CI=−1.44,-1.37; p<0.0001) and the MGB Biobank (ß=−1.25; SE=0.07; 95% CI=−1.38,-1.11; p<0.0001).

There was no significant difference in ANC between Black and non-Black participants with the rs2814778 CC variant in the UK Biobank (ß=0.05; SE=0.04; 95% CI=−0.02,0.13; p=0.172) or the MGB Biobank (ß=0.11; SE= 0.16, 95% CI=−0.22,0.43; p=0.517) nor any difference by age in the UK Biobank (ß=−0.002; SE=0.001; 95% CI=−0.005,0.003; p=0.086) or MGB Biobank (ß=−0.01; SE= 0.003, 95% CI=−0.013, 0.002; p=0.119). Female sex was significantly associated with higher ANC in the UK Biobank (ß=−0.19; SE=0.03; 95% CI=−0.24,-0.14; p<0.001) and the MGB Biobank (ß=−0.37; SE= 0.13, 95% CI=−0.62, −0.13; p=0.003). ANC data for the two biobank cohorts by Duffy status and racial category is shown in **Figure 4**.

**Figure 4:**
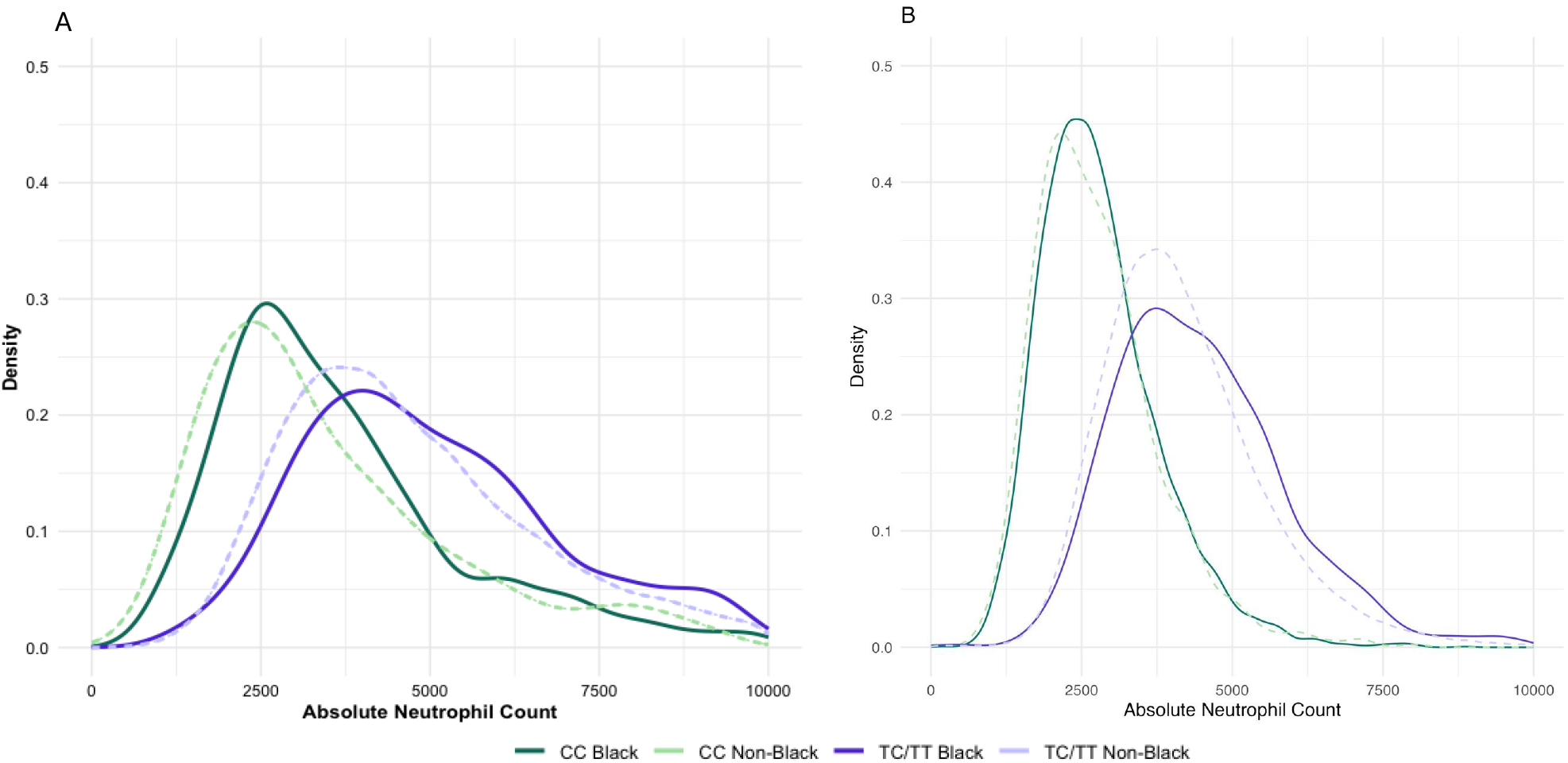
ANC Distribution by Duffy Genotype and Self-Reported Race. ANC values are presented for Duffy null (rs2814778 CC) and Duffy non-null (rs2814778 TC, CT, or TT) participants, stratified by self-identified Black and non-Black race. a) In the UK biobank, there is a significant difference in ANC by Duffy genotype (ß=−1.41; SE=0.02; 95% CI=−1.44,-1.37; p<0.0001), but no significant difference by self-identified race (ß=0.05; SE=0.04; 95% CI=−0.02,0.13; p=0.172). b) In the MGB Biobank, there is a significant difference in ANC by Duffy genotype (ß=−1.25; SE=0.07; 95% CI=−1.38,-1.11; p<0.0001), but no significant difference by self-identified race (ß=0.11; SE= 0.16, 95% CI=−0.22,0.43; p=0.517).

### White Blood Cell Counts

WBC for participants with the Duffy null phenotype in dedicated cohorts from Namibia, Saudi Arabia, and the USA is shown in **Table 1** and **Figure 3**. The nonparametric central two-sided 95% WBC interval indicating the 2.5% and 97.5% values was 2.51–9.85× 10^9^/L in Namibia, 3.71–9.95× 10^9^/L in Saudi Arabia, and 3.00-9.66× 10^9^/L in the USA. There was an overlap of 95% confidence intervals surrounding the upper and lower limits of normal between all three dedicated cohorts. Bootstrapped analysis of the three dedicated cohorts combined gives a 2.5% and 97.5% WCC value of 3.14 (95% CI: 2.85-3.49) and 9.80 (95% CI: 9.38-10.44), respectively.

WBC distributions differ across the three dedicated cohorts (p=0.002). There was no significant difference in WBC medians between the USA and Namibia (p = 0.561). There was a difference in WBC medians between Namibia and Saudi Arabia (p = 0.005) and between the USA and Saudi Arabia (p = 0.011). When only male data is analyzed, there is still a difference in WBC medians between Saudi Arabia and the USA (p=0.001) and Namibia and Saudi Arabia (p<0.001).

We also assessed the proportion of participants with a WBC below their respective institutional lower limit of normal (4.00× 10^9^/L in Namibia; 4.50× 10^9^/L in Jazan, Saudi Arabia; 4.00× 10^9^/L in Boston, USA). In total, 18% (n=22) of the Namibia cohort, 12.7% (n=19) of the Saudi Arabia cohort, and 10% (n=12) of the USA cohort had a WBC below their respective institutional reference interval.

## DISCUSSION

This multinational effort integrates biobank data with directly recruited cohorts of healthy participants and blood donors to investigate ANC distributions in Duffy null individuals across different continents and self-identified racial or ethnic groups. Additionally, we report the first WBC reference intervals for Duffy null individuals, which are significantly lower than current institutional reference intervals. While ANC and WBC distributions varied across cohorts, there were no significant differences in the upper or lower limits of normal.

Our data from Africa, Asia, Europe and North America emphasizes the influence of Duffy status on ANC and WBC reference intervals. In the dedicated cohorts, 21.7–50.7% of Duffy null participants had ANC values below their respective institutional reference intervals, with a similar but less pronounced pattern for WBC. Misclassifying Duffy null individuals as “abnormal” based on non-representative ANC and WBC reference intervals can lead to unnecessary interventions (e.g. bone marrow biopsies)^11,21^, over-investigation^22^, dose reductions for critical medications such as chemotherapeutics^23^ and clozapine^24^, and exclusion from clinical trials^25,26^. To avoid these inequities, we strongly recommend the development and adoption of separate ANC and WBC reference intervals for Duffy null and Duffy non-null individuals.

In exploring the consistency of Duffy null reference intervals across populations, we found some variation in distribution, but no significant differences in the upper and lower limits of normal. Reference limits are of primary importance for clinicians, as they define the boundary between normal and abnormal results. Differences in ANC medians between the Saudi Arabian and USA cohorts were explained by sex distribution; the Saudi cohort was 98.7% male, whereas the USA cohort was 26.7% male. When sex was accounted for, significant differences in ANC medians disappeared, consistent with known sex-related variations in ANC^22^. Persistent differences in median WBC, however, may reflect other unaccounted factors like lymphocyte count, tobacco exposure, or obesity^27,28^.

This data strongly challenges the use of race or ethnicity categories as proxies for biological differences. Race and ethnicity are social constructs rather than biological realities, with more genetic variability within racialized groups than between them^29^. In both the MGB Biobank and the UK Biobank, ANC did not differ significantly by racial or ethnic identity among Duffy null individuals. Furthermore, a significant number of Duffy null biobank participants identified with non-Black categories (12.1% in UK Biobank; 16.7% in the MGB Biobank). Conversely, a substantial proportion of individuals identifying as Black were Duffy non-null (35.8% in the MGB Biobank).^30^ These findings highlight the limitations of racial or ethnic identity as proxies and emphasize that Duffy status is the primary determinant of baseline ANC and WBC levels.

This study has several limitations. While dedicated cohorts met the CLSI minimum criterion of 120 for establishing new reference intervals, their sample sizes remain relatively small. Local laboratory practices introduce variability, and differences in sex distribution between cohorts impacted the ANC assessment. Dedicated cohorts excluded participants with conditions or treatments affecting neutrophil counts, but there are likely still additional unaccounted-for factors that can influence ANC and WBC. Additionally, MGB Biobank laboratory data is connected to the EHR and susceptible to collection during illness, likely explaining the higher ULN values compared to the UK Biobank or dedicated cohorts.

This study demonstrates that current institutional reference ranges disproportionately and inappropriately misclassify Duffy null individuals as neutropenic across all studied geographic, racial or ethnic backgrounds. This misclassification perpetuates systemic racism by privileging reference standards derived predominantly from individuals of European ancestry^22^. Many current ANC and WBC reference ranges do not adequately represent Duffy null individuals, predominantly harming racially marginalized groups within healthcare systems. Our findings highlight the inadequacy of current reference intervals for clinical decision-making and call on the World Health Organization (WHO), laboratories, and healthcare institutions worldwide to adopt Duffy-specific reference intervals to ensure informed and equitable healthcare for all patients.

## Author Contributions

SPH and LEM conceptualized the idea. IC, AJH, AAA, ES, MJA, HTC, MAN, JMS, MR, and LEM collected and curated the data. SPH, SEL, GF, DD, and LEM were involved in the formal analysis of the data. SPH and LEM acquired funding. SPH, IC, AJH, SEL, GF, MS, SP, and LEM contributed to the writing of the original draft. All authors contributed to reviewing and editing of the final version. SPH, IC, AJH, SEL, and LEM accessed and verified the underlying data reported in the manuscript. Coauthors are representative of the geography and racial/ethnic diversity discussed in this manuscript. All authors had full access to all the data in the study, reviewed the work, gave final approval of the version to be published, and agreed to be accountable for all aspects of the work.

## Data Sharing Statement

De-identified participant data and code used for statistical analysis and figure generation will be made available on written request to the corresponding author and after signed data access agreement after publication.

## Ethic Committee Approval

Ethics approval was obtained for all sites as outlined in the methods.

## Acknowledgement

The authors gratefully acknowledge Fanuel Mbago who assisted with sample analysis for Namibian participants, Natalia Simon who supported Namibian donor follow up, and Mass General Brigham Biobank for providing samples, genomic data, and health information data.

