## Supplementary Methods for "Multinational Assessment of Absolute Neutrophil Counts and White Blood Cell Counts Among Healthy Duffy Null Adults"

**Namibia**

Participants in this study must have met the eligibility criteria to be a blood donor. Exclusion criteria from the Blood Transfusion Service of Namibia are detailed in the “Guidelines for Medical Assessment of Blood Donors” (pages 4-172). In brief, these exclude individuals with cancer, infections, and most other serious comorbidities.

**Saudi Arabia**

Participants in this study must have met the eligibility criteria to be a blood donor. Exclusion criteria from Saudi Arabia Central Board for Accreditation of Healthcare Institution (CBAHI) Clinical Laboratories and Blood Banks Standards are as follows:

1. Whole blood is not collected from a donor weighing less than 50Kg or under 17 years of age.
2. Whole blood is not collected from a donor more frequently than once every eight weeks, not exceeding five times every twelve months and not from donors who donated apheresis product less than 48 hours ago.
3. The blood pressure and pulse rate of prospective donor are within normal ranges:
4. The hemoglobin level of the prospective donor should be greater than 12.5g/dL or a hematocrit of more than 38% for both male and female donors.
5. The prospective donor has no history of heart or lung disease
6. Female donors are not pregnant or have been pregnant within the last six weeks.
7. Prospective donor’s history is evaluated and the donor examined by qualified health care professional before whole blood collection.
8. Evidence of disease transmissible by blood transfusion.
9. Preventing donation by a person with other conditions thought to compromise the suitability of the blood or blood component.
10. The body temperature of the prospective donor not exceeding 37.5C.
11. The prospective donor has no history of liver diseases, cancer or bleeding tendency.
12. The prospective donor has no history of laboratory or clinical evidence for viral hepatitis, HIV, and HTLV.
13. The prospective donor has no history of laboratory or clinical evidence for malaria within the last three years.
14. The prospective donor has no history of blood transfusion or exposure to blood contaminated instruments (tattoo, cupping, or needle-stick injury) within the last twelve months.
15. The prospective donor has no history of syphilis treatment or unconfirmed test result for syphilis within the past 12 months.
16. The prospective donor has not been excluded as per the current recommendations for the prevention of HIV infection.
17. The prospective donor’s travel history checked against the current travel deferral list for the risk of HIV, vCJD and Malaria.
18. The prospective donor’s medications checked against current deferral list. Other medications are assessed by the blood bank physician.
19. The prospective donor’s vaccinations checked against the current vaccination deferral list. Other vaccinations must be assessed by the blood bank physician.
20. Prospective donor’s arms are free of lesions suggestive of skin disease or parenteral drug abuse.

**Black American Primary Care Patients (Boston)**

**eTable 1: Exclusion criteria for patients presenting for non-urgent care visits at a single primary care site**

| Autoimmune | Type 1 diabetes, rheumatoid arthritis, psoriasis, multiple sclerosis, systemic lupus erythematosus, inflammatory bowel disease, Addison’s disease, Graves’ disease, Hashimoto thyroiditis, Myasthenia gravis, Sjogren’s syndrome, pernicious anemia, autoimmune vasculitis, celiac disease, autoimmune hepatitis, vitiligo, immune thrombocytopenia, dermatomyositis. |
| --- | --- |
| Immunodeficiency | Any primary immunodeficiency including chronic granulomatous disease, common variable immunodeficiency, Chediak-Higashi syndrome, cyclic neutropenia, leukocyte adhesion defects, or congenital neutropenia. Any individual with previous assessment for or diagnosis of neutropenia of any kind was also excluded. |
| Infectious | Hepatitis C infection, hepatitis B infection, HIV |
| Malignancy | Any malignant neoplasm or hematological cancer including ductal carcinoma in situ, intraductal papillary mucinous neoplasm, or MGUS |
| Transplant | Any history of receiving organ transplant or skin graft including history of allogenic or autologous hematopoietic stem cell transplant |
| Medication | Carbimazole, clozapine, dapsone, dipyrone, methimazole, penicillin G, procainamide, propylthiouracil, rituximab, sulfasalazine, ticlopidine, hydroxychloroquine, infliximab, lamotrigine, oxacillin, quinine, infliximab, trimethoprim-sulfamethoxazole. Any chemotherapeutic agent or biologic |

**UK Biobank**

UK Biobank participants were genotyped on either the UK Biobank Axiom or the UK BiLEVE Axiom purpose-built arrays. Standard quality-control procedures were applied prior to imputation using the Haplotype Reference Consortium panel. A total of 487,323 individuals in UK Biobank had genotypes for rs2814778.

Absolute neutrophil count (field identifier: 30140) was measured at the initial assessment centre visit from blood samples. Neutrophil count is the proportion of (neutrophils / 100) x white blood cell count.

**Exclusions:** Participants with International Classification of Diseases 10^th^ Revision (ICD-10) diagnoses that are, or their treatment, associated with neutropenia were excluded. The non-cancer diagnoses excluded are detailed in eTable 2 and were defined from ICD-10 codes from hospital admissions (field identifiers 41202 and 41204) or death records (field identifiers 40001 and 40002), an equivalent read code from primary care records (field identifier 1712), or from a self-report questionnaire (field identifier 20002). The cancer ICD-10 diagnoses excluded using hospital admissions and death records included C00-C26, C30-C97 and C64-C48 inclusive. All cancer self-reported diagnoses (field identifier 20001) were excluded apart from non-melanoma skin cancer, basal cell carcinoma, and squamous cell carcinoma. A total of 141,228 participants were excluded with one or more diagnosis.

**eTable 2: Source of non-cancer diagnoses excluded from UK Biobank analyses**

| **Diagnosis** | **ICD-10 code** | **Codes included from field 20002 (self-report)** | **Codes included from field 1712 (GP records)** |
| --- | --- | --- | --- |
| Acute hepatitis B | B16 | 1579 | 130197 |
| Acute hepatitis C | B19 | 1580 | 130203 |
| Chronic viral hepatitis B with delta-agent | B16 |  |  |
| Chronic viral hepatitis B without delta-agent | B16 |  |  |
| Chronic viral hepatitis C | B18 |  | 130201 |
| Unspecified viral hepatitis B | B16 |  |  |
| Unspecified viral hepatitis C | B18 |  |  |
| Human immunodeficiency virus HIV disease | B20-B24 | 1439 | 130205; 130207; 130209; 130211; 130213 |
| Vitamin B12 deficiency due to intrinsic factor deficiency | D51.0 | 1331 |  |
| Congential agranulocytosis | D70 | 1448 | 130661 |
| Other drug induced agranulocytosis | D70 |  |  |
| Cyclic neutropenia | D70 |  |  |
| Functional disorders of polymorphonuclear neutrophils | D71 |  | 130663 |
| Immunodeficiency with predominantly antibody defects | D80 |  | 130677 |
| Combined immunodeficiencies | D81 |  | 130679 |
| Immunodeficiency associated with other major defects | D82 |  | 130681 |
| Common variable immunodeficiency | D83 |  | 130683 |
| Other immunodeficiencies | D84 |  | 130685 |
| Graft-versus-host disease | T86 |  |  |
| Thyrotoxicosis with diffuse goiter | E05.0 | 1522 | 130701 |
| Other thyrotoxicosis | E05.8 |  |  |
| Thyrotoxicosis, unspecified | E05.9 |  |  |
| Autoimmune thyroiditis | E06.3 |  |  |
| Type 1 diabetes mellitus | E10 | 1222 | 130707 |
| Primary adrenocortical insufficiency | E27.1 | 1234 | 130735 |
| Addisonian crisis | E27.2 |  |  |
| Other and unspecified adrenocortical insufficiency | E27.4 |  |  |
| Multiple sclerosis | G35 | 1261 | 131043 |
| Myasthenia gravis and other myoneural disorders | G70 | 1260; 1437 | 131093 |
| Crohn's disease | K50 | 1462 | 131627 |
| Ulcerative colitis | K51 | 1463 | 131629 |
| Autoimmune hepatitis | K75.4 |  |  |
| Celiac disease | K90.0 |  |  |
| Psoriasis | L40 | 1453 | 131743 |
| Parapsoriasis | L41 |  | 131745 |
| Discoid lupus erythematosus | L93 | 1381 | 131829 |
| Vasculitis limited to the skin, not elsewhere classified | L95 |  | 131833 |
| Rheumatoid arthritis with rheumatoid factor | M05 | 1464 | 131849 |
| Other rheumatoid arthritis | M06 |  | 131851 |
| Psoriatic and enteropathic arthropathies | M07 |  | 131853 |
| Juvenile RA, AS | M08 |  | 131853 |
| Polyarteritis nodosa | M30.0 | 1380 | 131891 |
| Churg-Strauss | M30.1 |  |  |
| Mucocutaneous lymph node syndrome | M30.3 |  |  |
| Wegener's granulomatosis | M31.3 | 1378 | 131893 |
| Aortic arch syndrome | M31.4 |  |  |
| Giant cell arteritis with polymalgia rheumatica | M31.5 |  |  |
| Other giant cell arteritis | M31.6 | 1376 |  |
| Microscopic polyangiitis | M31.7 | 1379 |  |
| Systemic lupus erythematosis with organ or system involvement | M32.1 | 1381 | 131895 |
| Other forms of systemic lupus erythematosus | M32.8 |  |  |
| Systemic lupus erythematosis, unspecified, multiple sites | M32.90 |  |  |
| Systemic sclerosis | M34 | 1384 | 131899 |
| Dermatopolymyositis | M33 | 1383 | 131897 |
| Sjogren syndrome | M35.0 | 1382 | 131901 |
| Polymyalgia rheumatica | M35.3 | 1377 |  |
| Ankylosing spondylitis | M45 | 1313 | 131913 |
| Bechet disease | M35.2 |  |  |

**MGB Biobank**

The MGB Biobank (formerly named Partners Biobank) is an ongoing observational research project that enrolls patients and employees of a multicenter health system in Eastern Massachusetts. Participants are enrolled using a broad-based consent process by up to 30 research coordinators located at health system practices, in public hospital locations, as part of a collaborating study (dual consent) or electronically through Patient Gateway, the MGB patient portal. Demographic data and blood samples are collected at baseline and linked to EHRs data and self-reported health surveys for ongoing research.

Biobank samples are genotyped using 3 versions of the Multi-Ethnic Global BeadChip SNP array offered by Illumina that is designed to capture the diversity of genetic backgrounds. These arrays cover over 1.7 million unphased variants which are annotated for dbSNP rs identifier, gene location, and protein and variant effect using Alamut-Batch (Interactive Biosoftware, France, <https://www.interactive-biosoftware.com/alamut-batch/>). Genotype calls and annotations are made available to investigators as VCF and PED file formats. Imputed genotypes are also available. All patients with rs2814778 were selected for. Those with the CC variant are Duffy null and those with the CT, TC, or TT variant are Duffy non-null.

Computed phenotypes are derived from both structured and unstructured EHR data and provide the ability for researchers to accurately select a disease population for genomic or other analyses. The Biobank Portal computed phenotypes (also called “Curated Disease Populations”) are trained using PheCAP, a well-defined supervised learning workflow. Once a model is trained, it is operationalized in the data repository build process using SQL scripts that define features based on ontology paths and run the prediction to estimate predicted probability of each patient having a disease.

As trained phenotypes are more accurate than individual ICD-10 codes, these were used to identify and exclude patients with diseases that are likely to impact ANC. Phenotypes are listed in a binary form (yes/no). Phenotypes assessed for and excluded from the cohort include HIV infection, systemic autoimmune disease, rheumatoid arthritis, noninfective inflammatory bowel disease, celiac disease, hepatitis C antibody, chronic viral hepatitis C, myeloproliferative diseases, and personal history of malignant neoplasms of other organs and systems.
